# Returning to a normal life via COVID-19 vaccines in the USA: a large-scale agent-based simulation study

**DOI:** 10.1101/2021.01.31.21250872

**Authors:** Junjiang Li, Philippe J. Giabbanelli

**Affiliations:** Department of Computer Science & Software Engineering, Miami University, Oxford OH, 45056 USA

**Keywords:** Agent-Based Model, Cloud-Based Simulations, COVID-19, Large-Scale Simulations, Vaccine

## Abstract

**Background:** In 2020, COVID-19 has claimed more than 300,000 deaths in the US alone. While non-pharmaceutical interventions were implemented by federal and state governments in the USA, these efforts have failed to contain the virus. Following the FDA approval of two COVID-19 vaccines, however, the hope for the return to normalcy is renewed. This hope rests on an unprecedented nation-wide vaccine campaign, which faces many logistical challenges and is also contingent on several factors whose values are currently unknown.

**Objective:** We study the effectiveness of a nation-wide vaccine campaign in response to different vaccine efficacies, the willingness of the population to be vaccinated, and the daily vaccine capacity under two different federal plans. To characterize the possible outcomes most accurately, we also account for the interactions between non-pharmaceutical interventions and vaccines, through six scenarios that capture a range of possible impact from non-pharmaceutical interventions.

**Methods:** We use large-scale cloud-based agent-based simulations by implementing the vaccination campaign using Covasim, an open-source ABM for COVID-19 that has been used in several peer-reviewed studies and accounts for individual heterogeneity as well as a multiplicity of contact networks. Several modifications to the parameters and simulation logic were made to better align the model with current evidence. We chose six non-pharmaceutical intervention scenarios and applied the vaccination intervention following both the plan proposed by Operation Warp Speed (former Trump administration) and the plan of one million vaccines per day, proposed by the Biden administration. We accounted for unknowns in vaccine efficacies and levels of population compliance by varying both parameters. For each experiment, the cumulative infection growth is fitted to a logistic growth model, and the carrying capacities and the growth rates are recorded.

**Results:** For both vaccination plans and all non-pharmaceutical intervention scenarios, the presence of the vaccine intervention considerably lowers the total number of infections when life returns to normal, even when the population compliance to vaccines is as low at 20%. We noted an unintended consequence: given the vaccine availability estimates under both federal plans and the focus on vaccinating individuals by age categories, a significant reduction in non-pharmaceutical interventions results in a counterintuitive situation in which higher vaccine compliance then leads to more total infections.

**Conclusions:** Although potent, vaccines alone cannot effectively end the pandemic given the current availability estimates and the adopted vaccination strategy. Non-pharmaceutical interventions need to continue and be enforced to ensure high compliance, so that the rate of immunity established by vaccination outpaces that induced by infections.

## Introduction

The Centers for Disease Control and Prevention (CDC) forecasted that 300,000 deaths would be attributable to COVID-19 by the end of the year. Reality defied expectations, as COVID-19 was *directly* responsible for approximately 350,000 deaths in the USA out of 20 million *reported* cases (for forecasts and total case numbers, see [1]), which may only represent one out of seven actual cases based on CDC estimates for September [2]. Despite popular comparison with the flu, the ongoing COVID-19 epidemic has thus already claimed five times as many lives than the worst year for the flu, whose recent yearly death tolls range from a low of 16,000 to a high of 68,000 [3]. To contextualize the impact of COVID-19, we note that the U.S. *life expectancy* decreased by more than a year, which is ten times worst than the decline from the opioid epidemic [4]. In another comparison, 2020 is the *largest single-year increase in mortality* in the USA since 1918, which had both a flu pandemic and a war. This reflects both direct and *indirect* consequences on COVID-19, such as disrupting in-person treatments [5] and supply networks, with effects as far ranging as a jump in drug overdose [6]. To complement measures of short-term effects such as deaths or number of cases, we also note the long-term impacts captured by the outpatient journey. Common symptoms often persist over month (e.g. fatigue, cough, headache, sore throat, loss of smell) [7–9] and less frequent ones can be severe since COVID-19 involves many organs. Effects can involve the cardiovascular system in up to 20-30% of hospitalized patients [10–11] (e.g., cardiac injury, vascular dysfunction, thrombosis), result in kidney injury [10] or pulmonary abnormalities [13], or lead to a deterioration in cognition due to cerebral micro-structural changes [14]. Based on similar infections, such effects can be long: for instance, inflammation of the heart caused by viral infections (i.e., myocarditis) can have a recovery period spanning months to years.

Interventions in 2020 were strictly *non-pharmaceutical*, as vaccines were being developed and tested. Such intervention strategies have included preventative care (e.g., social distancing, hand washing, face masks), lockdowns (e.g., trasvel restrictions, school closures, remote work), and logistics associated with testing (e.g., contact tracing, quarantine) [15–16]. The range of non-pharmaceutical interventions adopted at various times across countries can be seen in further details through the CoronaNet project [17] or the collection of essays “mobilizing policy (in)capacity to fight COVID-19” published in mid-2020 [18]. In early 2021, two vaccines were deployed (Pfizer-BioNTech and Moderna) with plans for up to three additional vaccines (AstraZeneca, Janssen, Novavax) [19]. With the availability of vaccines comes the key question: when will life return to normal in the USA? The implicit expectation is to see a return to normalcy thanks to the vaccine, rather than due to a high number of cases with its accompanying death toll.

In a highly publicized interview, Dr. Anthony Fauci, director of the National Institute of Allergy and Infectious Diseases, estimated a return to normal by fall, *if the vaccination campaign is successful* [20]. Getting a precise estimate of when life will return to normal is a challenge as it depends on numerous interrelated factors: potential behavioral changes affecting non-pharmaceutical approaches (e.g., lesser compliance to mask wearing and social distancing), participation in the vaccination campaign, logistics associated with vaccination (i.e., who can get vaccinated and when), and mutations leading to new strains with different biological properties (e.g., higher infectivity) or unknown vaccine responses. In this paper, we use large-scale simulations to identify *when* there will be an inflection point in the dynamics of the disease, and the *level* of cases that will be obtained.

Simulations have been used since the early days of the COVID-19 pandemic. Classic compartmental epidemiological models were first produced (e.g., many SEIR models [21–24]), with a focus on estimating broad trends and key epidemiological quantities such as the expected number of new cases generated by each infected individual (i.e., the basic reproduction number R_0_). Such compartmental models provide limited support to study the effect of interventions, for instance by lowering the contact rate to represent the impact of social distancing. A research shift in the second part of 2020 resulted in the growing use of *Agent-Based Models* (*ABM*) to support the analysis of interventions by explicitly modeling each individual as well as their interactions among each other or with the environment. This shift to individual-level models was underpinned by the evidence of *heterogeneity* in risk factors (e.g., older age, hypertension, respiratory disease, cardiovascular disease [25–26]) as well as behaviors (e.g., non-compliance with social distancing orders) based on personal beliefs and values [27–28]. There is also spatial variation in socio-ecological vulnerability to COVID-19 [29], with rural counties being at higher risk (due to e.g., older population with more underlying conditions, lower access to resources) [30–31] and hence experiencing higher mortality rates [32]. Finally, there is a documented heterogeneity in transmission based on contact tracing data [33], which stresses the need to use realistic networks when modeling the spread of COVID-19 [34]. Considering this growing evidence-base, our work relies on an ABM which accounts for individual heterogeneity (e.g., in age), explicitly embeds them in a network to model their contacts, and simultaneously considers different network types (e.g., community, work) to account for various settings.

By adding vaccines to a previously validated ABM of COVID-19, we are able to assess how the number and timing of cases depends on key factors such as the population’s interest in vaccines and the efficacy of vaccines. Our specific contributions are twofold:

- We extend the validated COVASIM model with a detailed process of vaccination, accounting for vaccine efficacy, interest in vaccination, and fluctuations in vaccination capacity. Our process models the need for two doses and the possibility of being infected until the second dose is administered.
- We examine vaccination interventions under two hypotheses for the number of doses available and considering concurrent non-pharmaceutical interventions.

The remainder of this paper is structured as follows. In our methods, we briefly cover the rationale for choosing COVASIM and how we adapted the model to account for the latest epidemiological evidence. We then explain which non-pharmaceutical interventions are simulated, in line with our previous work [35]. Most importantly, we detail the novel extension of vaccines into COVASIM and our examination of the trends in cumulative infections using a logistic growth model. The following section presents and analyzes our results. Our final section discusses our main findings and provides an exhaustive list of limitations due to the ongoing nature of the pandemic and challenges in vaccination.

## Methods

### Overview

COVASIM was developed under leadership of the Institute for Disease Modeling and released in May 2020 by Kerr and colleagues [36]. It is one of several open-source Agent-Based Models, together with OpenABM-Covid19 [37] or COMOKIT [38]. The model captures the transition from susceptible to infected followed by a split between asymptomatic individuals and various degrees of symptoms, resulting either in recovery or death (Figure 1). The model was created to support interventions offered at the time, which did not include vaccination. We thus *modified the model* to account for our current understanding of viral dynamics as well as the use of vaccines over two doses (Figure 1). When instantiating the model to the U.S. population, we use a resolution of 1:500 (i.e., each simulated agent accounts for 500 U.S. inhabitants). Given our resolution and target population size, our application exceeds half a million agents and can thus be described as a “*large-scale COVID-19 simulation*” [39]. Our simulations start on January 1^st^ using CDC data for the number of infected, recovered, and immunized individuals to date (see subsection “Initializing the model”). We then simulate for 6 months, that is, 180 time ticks based on a temporal resolution of one day per simulation step (i.e., ‘tick’). To cope with the computational challenges created by a large-scale stochastic model, a philanthropic grant supports us in performing cloud-based simulations via the Microsoft Azure platform.

**Figure 1.**
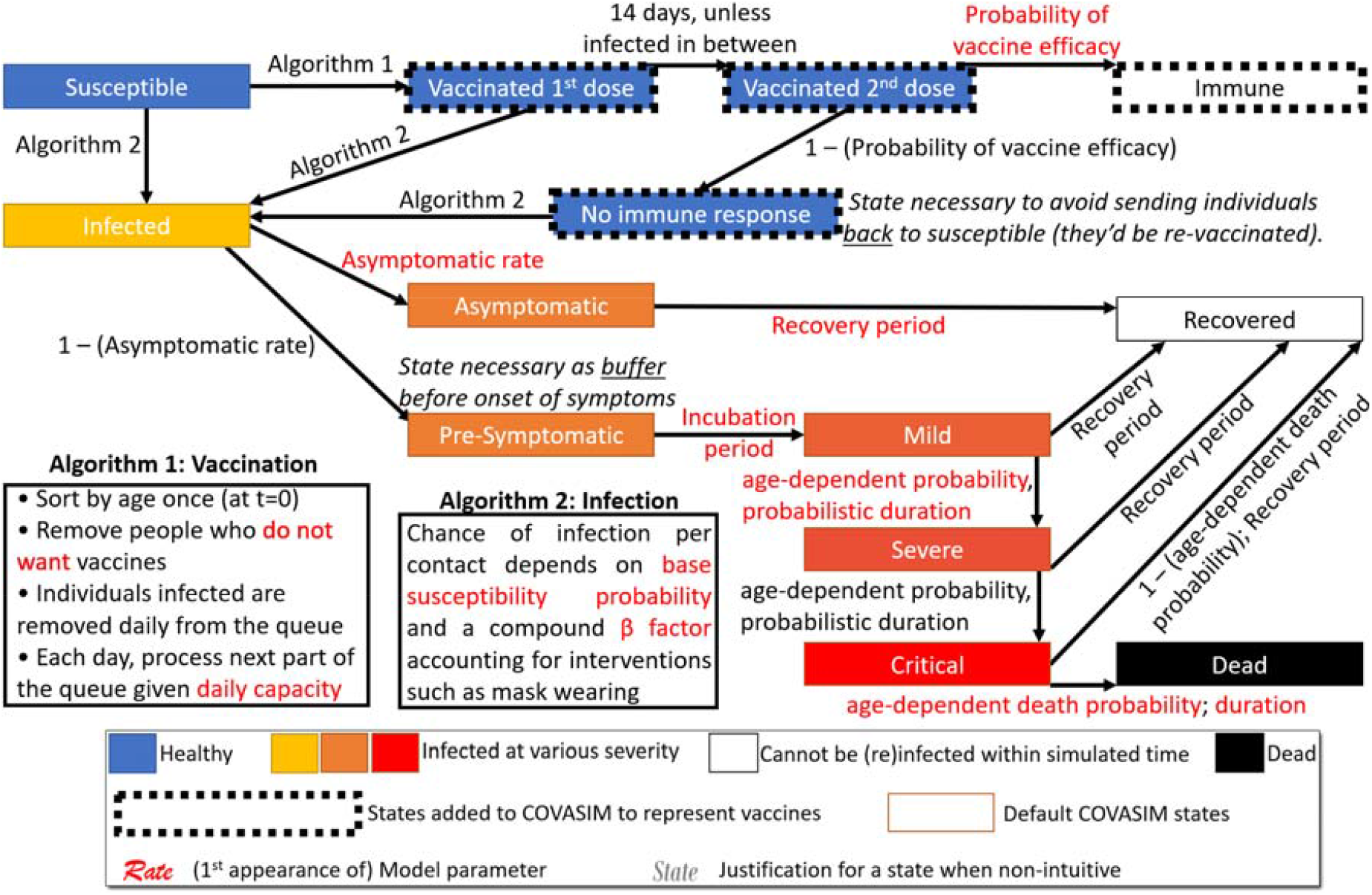
Overview of our modified COVASIM model containing the state diagram and specification of all transitions, including key procedures for vaccination and infection.

### The COVASIM model: rationale for selection and evidence-based updates

Apart from being open source, there are two reasons for which we selected COVASIM. First, it captures heterogeneity within individuals (e.g., assigns an age and uses age-specific disease outcomes) as well as transmission patterns, by placing agents within synthetic networks corresponding to a multiplicity of contexts: work (based on employment rates), school (based on enrollment), home (based on household size), and the general community. However, these high-resolution age-specific contact patterns are not unique to COVASIM. For example, the OpenABM-Covid19 [37] also embeds agents in age-stratified occupation networks (encompassing work and school), household networks, and a ‘general’ random network. COMOKIT [38] similarly uses the Gen* toolkit from the same team to redistribute populations from census units down to exact buildings such as the nearest school. Thus, the second rationale for choosing this platform is that it has been used in the most peer-reviewed modeling studies to date [40–41], hence providing an additional layer of scrutiny and confidence in the correctness of the model (i.e. validation) as well as its implementation (i.e. verification). As detailed in our recent work [35], changes in the evidence-base have required alteration in the model to keep it valid. Consequently, we modified three COVASIM parameters to account for the current biological and epidemiological evidence on COVID-19 (Table 1).

**Table 1.** Adjusted parameters based on reports in the U.S.

| COVASIM Construct | Initial value | Modified value | Rationale for modification |
| --- | --- | --- | --- |
| Incubation: delay from infection to viral shedding | Lognormal(4.6, 4.8) | Lognormal(4.1, 4.8) | The combined distribution of incubation period did not match latest evidence. The adjustment aligns it with the evidence. |
| Incubation: delay from viral shedding to onset of symptoms | Lognormal(1,1) | Lognormal(1, 1.8) |  |
| Proportion of symptomatic cases | 0.7 | 0.6 | Although reports vary, Dr Fauci stated that 40% of the US cases were asymptomatic. |

### Selection and representation of concurrent non-pharmaceutical interventions

In addition to support for heterogeneity, COVASIM implements several non-pharmaceutical interventions. Although our focus is on vaccines, such interventions may be continuing in parallel with the vaccination campaign hence we have to take them into account when forecasting case counts. Interventions can be organized into three broad categories: preventative care (e.g., *social distancing* and *face masks*), lockdown (e.g., *stay-at-home* orders such as remote work and/or school closures), or testing-related (e.g., *testing* itself, then *quarantining* and *contact tracing*) [42–43, 15]. In line with our previous work on non-pharmaceutical interventions, we considered all six specific interventions. Although all six are natively supported by the COVASIM platform, we changed testing delays from their default value (constant) to a distribution (based on a survey across all 50 U.S. states) [44], thus accounting for the variability observed in practice.

Since our focus is on vaccines, our search space is primarily devoted to quantifying the effect of vaccine-related variables (i.e., efficacy, compliance, capacity). As every non-pharmaceutical intervention could lead to a several variables (e.g., compliance with face masks, efficacy of face masks), considering all variables for every such intervention *in addition to* vaccine-related variables would lead to an impractical search space. We thus leveraged the systematic assessment of our previous study [35], which simulated all combinations of non-pharmaceutical interventions at two different levels of strength (i.e., a binary factorial design of experiments). We analyzed results from this broad search to select five scenarios (Table 2) that result in five different levels of infection count after six months, in the absence of any vaccine (Figure 2). In other words, to circumvent the unwieldy notion of simulating all aspects of vaccines and non-pharmaceutical interventions, we selected five scenarios that produce linear to logistic growths in cumulative infections, thereby conducted a parameter sweep across possible growth behaviors. We supplemented these five scenarios with an extreme “no intervention” scenario, which provides an upper bound on the number of cases.

**Table 2.** Scenarios depicting concurrent non-pharmaceutical interventions, chosen for their ability to create five markedly different outcomes together with a no-intervention case.

| Scenario →<br>Features ↓ | 1 | 2 | 3 | 4 | 5 | 6 (do nothing) |
| --- | --- | --- | --- | --- | --- | --- |
| Networks impacted | Work, school |  | Community |  |  | All |
| Percentage of contact in work and school (as a function of default) | 70% | 95% | N/A |  |  | 100% |
| Percentage of contact in community (as | N/A |  | 70% | 70% | 90% | 100% |
| a function of default) |  |  |  |  |  |  |
| Daily tests | 1.11M | 600K | 600K | 1.11M | 600K | No testing |
| A positive test leads to quarantine. Is a 2<sup>nd</sup> test required to end quarantine? | No | Yes | No | Yes | Yes |  |
| Test sensitivity | 1 | 1 | 1 | 0.55 | 0.55 |  |
| % of contacts that can be traced | 0.2 | 1 | 1 | 0.2 | 0.2 | No tracing |
| After how many days will contact tracing results arrive? (i.e. contact tracing delay) | 0 | 7 | 7 | 7 | 7 |  |
| Starting contact tracing if one has just been tested and exposed (one infected peer) | Yes | No | No | Yes | No |  |

**Figure 2.**
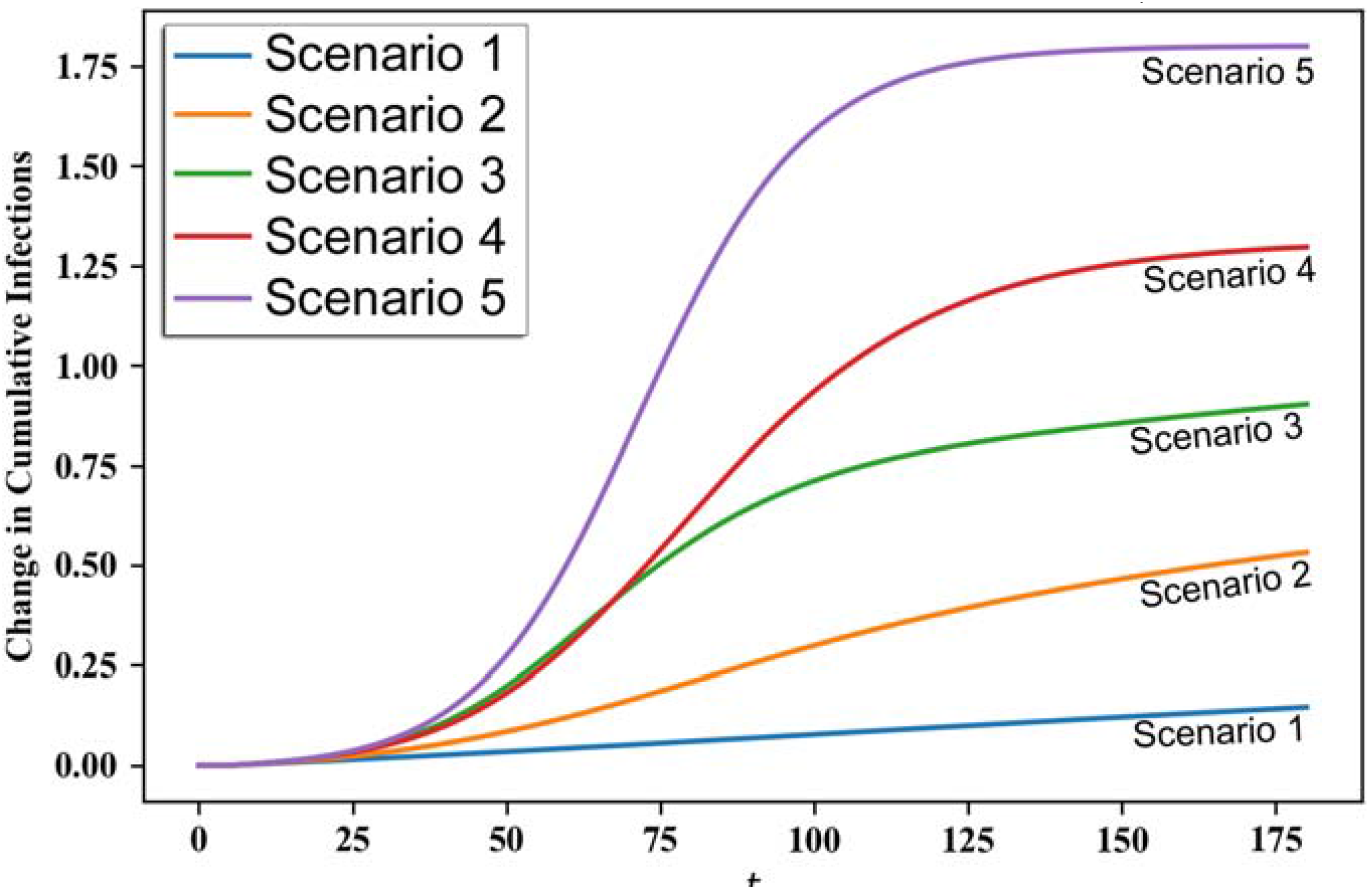
Five scenarios (each based on a combination of interventions), selected for their ability to represent different trends in the number of cases over time, without a vaccine.

Given that we made minor changes to the biology (incubation and proportion of symptomatic cases) and consider several ongoing intervention scenarios, it is necessary to confirm the validity of the model established using earlier data in previously published studies. Consequently, we ran the modified COVASIM model based on data observed until 9/3/2020 and compared the simulated results with observations until the end of year. Similar trends and orders of magnitude were observed (Figure 3), thus providing qualitative validation. Note that the five scenarios chosen (Table 2) bound the growth of COVID-19 in the US, such that we are comprehensively examining possible trends going forward instead of limiting ourselves to the single trend that fit best on previous data.

**Figure 3.**
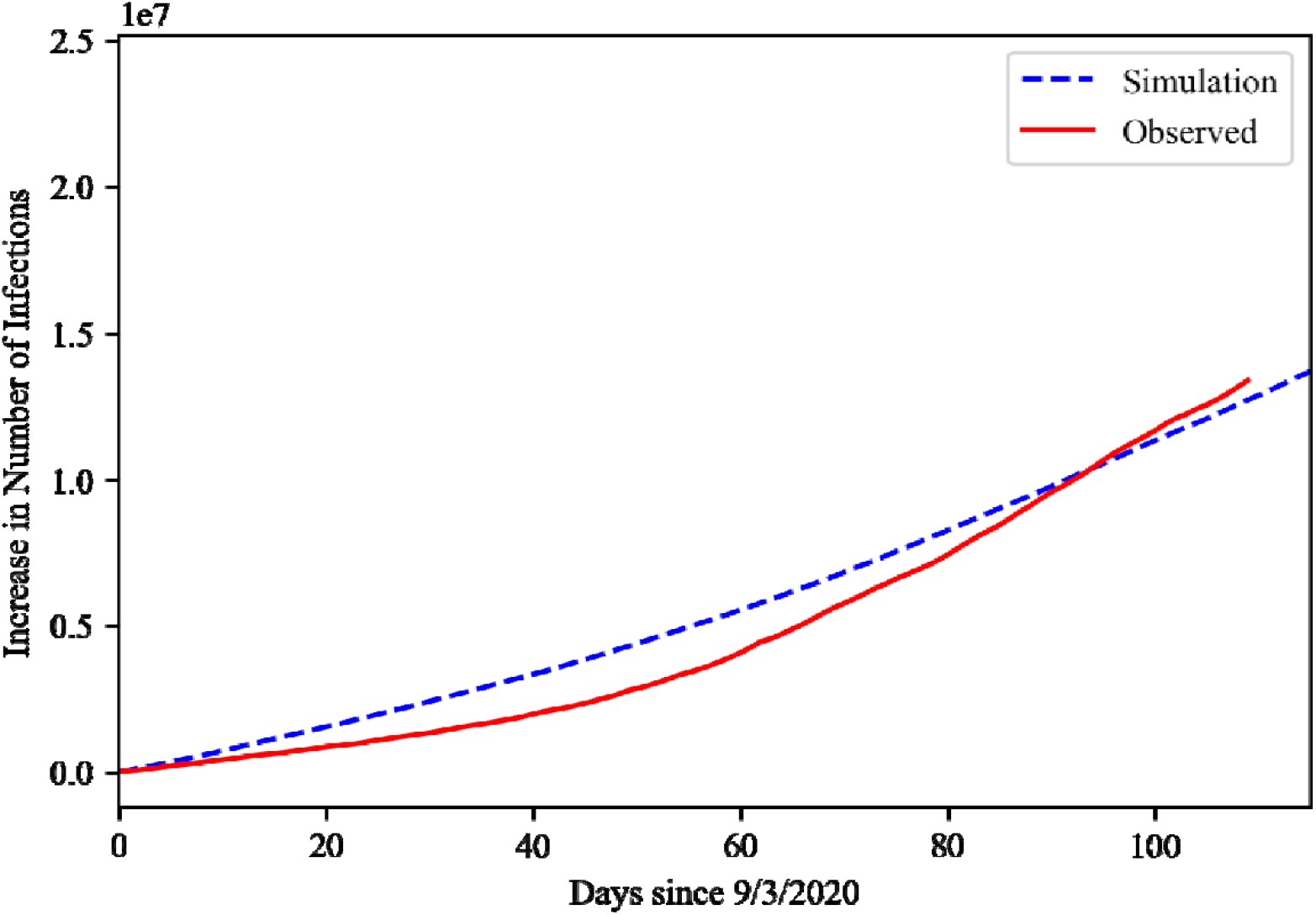
Comparison of changes in cumulative infections between a Covasim simulation and reality from 9/3/2020 to the end of 2020. The simulation included a reduction on work and school contacts (set to 95% of their capacity), 1.11M daily and highly sensitive tests, quarantining upon testing, immediate contact tracing with a level of 0.2, and a presumptive approach.

### Extending COVASIM with pharmaceutical interventions: a two-steps vaccination

As detailed in our discussion, there is significant uncertainty and frequent changes regarding the number of vaccines that *may* be administered monthly. We thus consider two vaccine availability scenarios, both proposed by federal governments. The first scenario from the former Trump administration, named Operation Warp Speed, stated that vaccines will be available in tiered amounts (20 million in December, 30 million in January, and 50 million every month thereafter). The second scenario from the Biden administration, known as the ‘100-day goal’, proposes that there will be 1 million vaccines every day [45], thus covering 50 million Americans. Although there are other scenarios, they vary from state to state (e.g., the governor of New Jersey aspires to vaccinate 70% of the adult population within 6 months [46]) and are also subject to frequent revisions. Given the country-wide nature of our simulation, we rely on federal plans while detailing challenges (c.f. discussion).

In setting the monthly capacity, we noticed the necessity to *adjust the schedule* of the Operation Warp Speed plan, since the initial aim of 20 million people immunized by the end of December only resulted in 3 million doses administered. In other words, it would be incorrect to model the monthly capacity of Operation Warp Speed as announced since there is evidence that its initial objective was unmet, due to a variety of logistical challenges [47]. Consequently, we shifted the expectations of the Operation Warp Speed plan by one month, such that the capacity for January now corresponds to the initial expectations for December (20 million) and so forth.

At the same time as either vaccination schedule is active, we also have the six scenarios listed in the previous sections. As these scenarios include a no-intervention case, we are able to study the interaction between non-pharmaceutical interventions and vaccines. In total, this gives 12 distinct situations. In addition, we also vary two essential parameters regarding vaccines: the percentage of the population that seeks vaccination (which we refer to as ‘*vaccine compliance*’ from hereon) and the efficacy of the vaccine. Varying these two parameters across 12 situations in a large-scale ABM results in significant computing needs. These are challenging to parallelize as the runtime of each experiment is not the same. Therefore, we took advantage of the massive parallelism enabled by the cloud computing platform Azure to accelerate computation. Using this platform, we varied vaccine compliance and vaccine efficacy between the bounds listed in Table 3.

**Table 3.** Vaccine parameters used in the study. Intermediate values in the interval bounded by the low and high values are automatically explored.

|  | Low Value | High Value |
| --- | --- | --- |
| Vaccine Compliance | 20% | 60% |
| Vaccine Efficacy | 88% | 99% |

Regarding our approach to vaccine efficacy, we note that individuals can be infected after their first dose, as has been documented on thousands of cases [48]. We thus only apply the probability of vaccine efficacy only after the 2nd dose. Although we do not track which of the two approved mRNA COVID-19 vaccine (Pfizer-BioNTech or Moderna) is administered, we vary vaccine efficacy to account for a margin of uncertainty regarding their respective performances. Since the vaccine capacity is either planned to increase (Operation Warp Speed) or at a high constant rate, a simulated agent given one dose will always be able to come back to get the second dose on time. Should an agent be contaminated or die before the second dose, it is then released for administration to another agent.

We also vary the percentage of the population who seeks vaccination. As noted in a recent study, this percentage has varied among studies: 10.8% did not intend to be vaccinated when asked in April 2020, but this number jumped to 31.1% by May, and an August poll found that only a *minority* would want to be vaccinated [49]. In addition to changes in the sociopolitical climate and public discourse surrounding vaccination, there will also be changes since “many receptive participants preferred to wait until others have taken the vaccine” [50]. Seeing positive vaccination outcomes in others may in part address the fear of serious side effects, which is a recurring concern for individuals who may not intend to participate in vaccination [51]. Given past variations and changes in the future, we handled uncertainty through a parameter sweep in vaccine compliance.

### Initializing the model

A simulation model is composed of an initialization (setting characteristics of agents for t=0) and rules governing its update, thereby producing data for analysis. The previous subsections covered the rationale for inclusion of agents’ characteristics and the design of the rules, while the next subsection focuses on the analysis. The present subsection thus briefly covers our approach to initialization such that our results can be independently replicated by other modeling teams.

Our initial time tick t=0 corresponds to January 1^st^ 2020. We thus need to set the number of agents who have been infected, recovered, or immunized (due to the rollout of vaccines in December) by that time. A COVID-19 case remains infectious within a time window of two weeks, after which there is either recovery or complications. From December 18^th^ to 31^st^, there was a total of 3,311,345 active cases. To appropriately initialize our simulation, we need to further track *when* an individual was infected. Incorrectly setting them to be all infected on December 18^th^ would result in nobody being infected when the simulation starts on January 1^st^. At the other extreme, assuming that they were all infected on December 31^st^ would lead to an overestimate of disease spread into 2021. We thus seeded the timing of each infection by using the daily distribution from CDC data between December 18^th^ and 31^st^ (Table 4). All numbers were divided by 500 since our agent resolution is 1 agent for 500 real-world U.S. inhabitants (1:500). The number of individuals who acquired immunity via recovery was set to the total case count observed by December 17^th^. Individuals who died from COVID-19 are grouped together with recovered ones (i.e., we do not subtract them from the count) since our simulations track the number of new infections: dead individuals do not alter these results as they can neither be infected nor infect others. The total number of individuals immunized from vaccination was set to 2 million (i.e. 4,000 agents).

**Table 4.**
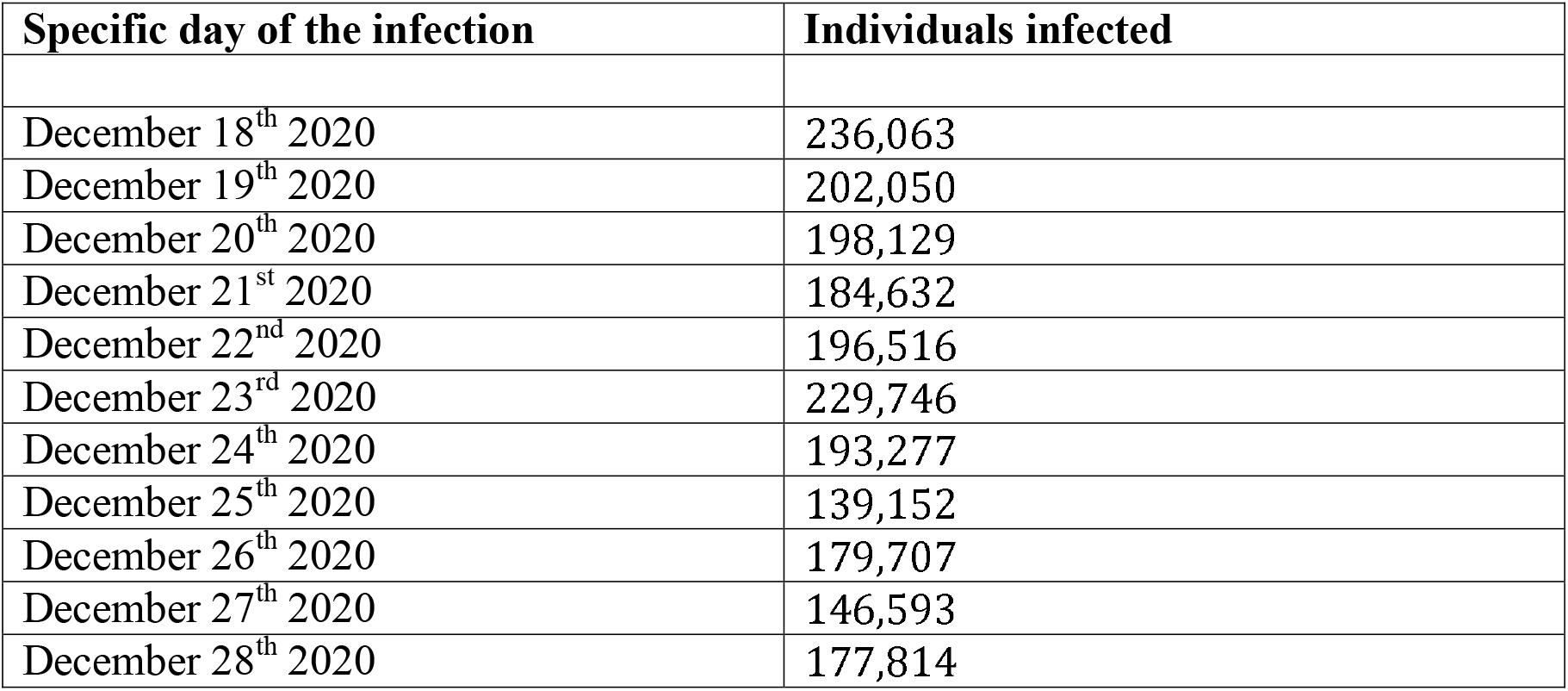
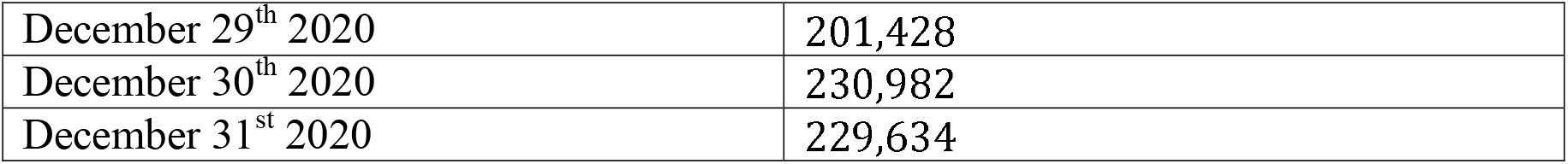
Timing of the infection in the two weeks preceding the start of our simulation, such that our agents can be initialized at the appropriate state of their infection.

### Analyzing the progression of cumulative infections through a logistic growth model

To quantify the spread of the disease, we fitted the progression of cumulative infection to a logistic growth model, which is a simple yet effective model describing resource-limited growths in natural processes and has been used on several occasions for COVID-19 [52–54]. Let the cumulative infection be *P*= *P*(*t*), then the logistic model stipulates that *P* is the solution of the differential equation

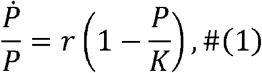

where 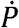 is the time derivative of *P, r* is the *growth rate* (proportional to the maximum value attained by 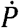), and *K* is the *carrying capacity*. As our simulations produce the complete time series for *P*, we can estimate 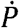 using finite differences, thereby extracting parameters *r* and *K* through a linear regression as equation (1) suggests. In the regression, the independent and dependent variables are *P* and 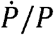, respectively. In addition, we measure the goodness of fit as that of the linear regression. Since the simulation is stochastic, multiple replications are needed for each configuration to obtain an average behavior. We used the confidence interval method [55, pp. 184-196] to perform enough replications so that for every time step *t*, the 95% CI of *P* at time *t* falls within 5% of the average. Therefore, we perform the fitting for each individual run, and compute the average *r* and *K* across all runs.

Although we report *r* and *K* in our supplementary material, the interpretation of these variables can be difficult for a broader audience. The growth rate *r* is *proportional* to the maximum *fraction of the carrying capacity K* that is infected on the worst day. In other words, it is an indication of how fast the disease spreads as its peak, based on another variable. For ease of interpretation, we focus on the adjusted growth rate whose unit is directly in number of individuals. This adjusted growth rate is obtained as

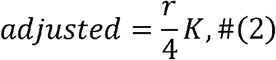

For instance, an adjusted value of 200,000 means that at most 200,000 individuals will be infected on the worst day.

As the early steps of the simulation witness a shift from a vaccine-naïve population to one that gradually builds vaccine-based immunity, early trends differ from the longer ones that are the focus of this study. This is a typical situation in modeling, whereby estimating the long run performance measures requires to first run the model for a certain amount of time (known as ‘warm-up period’) [56]. We empirically determined that a warm-up period of 20 days was sufficient to start the curve fitting, that is, we create the time series for *P* starting from *t* ≥20. As evidenced by Figure 4, this warm-up period results in very good fit for the logistic model under both federal plans. This approach also generalizes better, since the reported and can accurately characterize the spread of the disease for most time periods instead of being skewed by the first few days.

**Figure 4.**
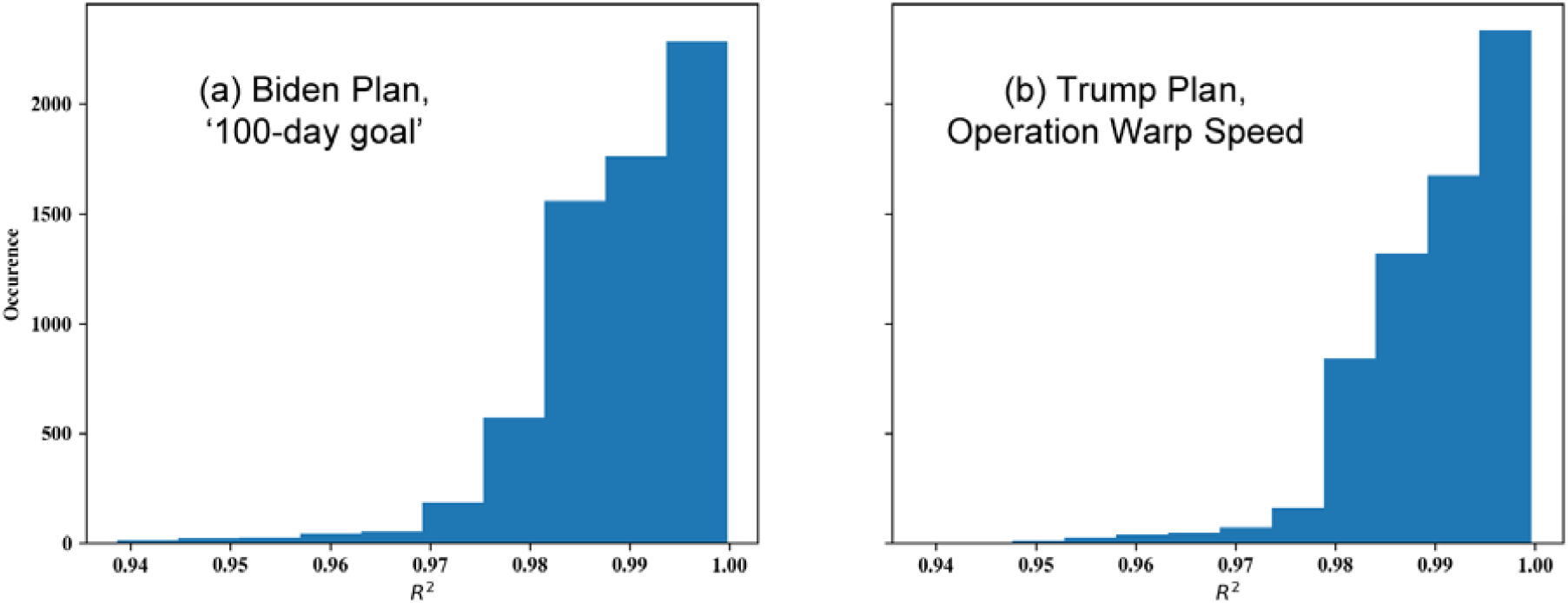
Distributions of the average goodness of fit for each vaccination plan, demonstrating the validity of fitting logistic growth models from.

An essential aspect of a return to normalcy is about the *conditions* under which that is achieved. If the disease is left uncontrolled, and simplifying the matter of variants, we would still return to ‘normalcy’ within six months because a very large share of the population would already have been infected and either recovered or died (Figure 5). The goal is thus not *only* to eventually achieve stability in the number of cases but to achieve it at a minimal level (Figure 5; bottom blue curve).

**Figure 5.**
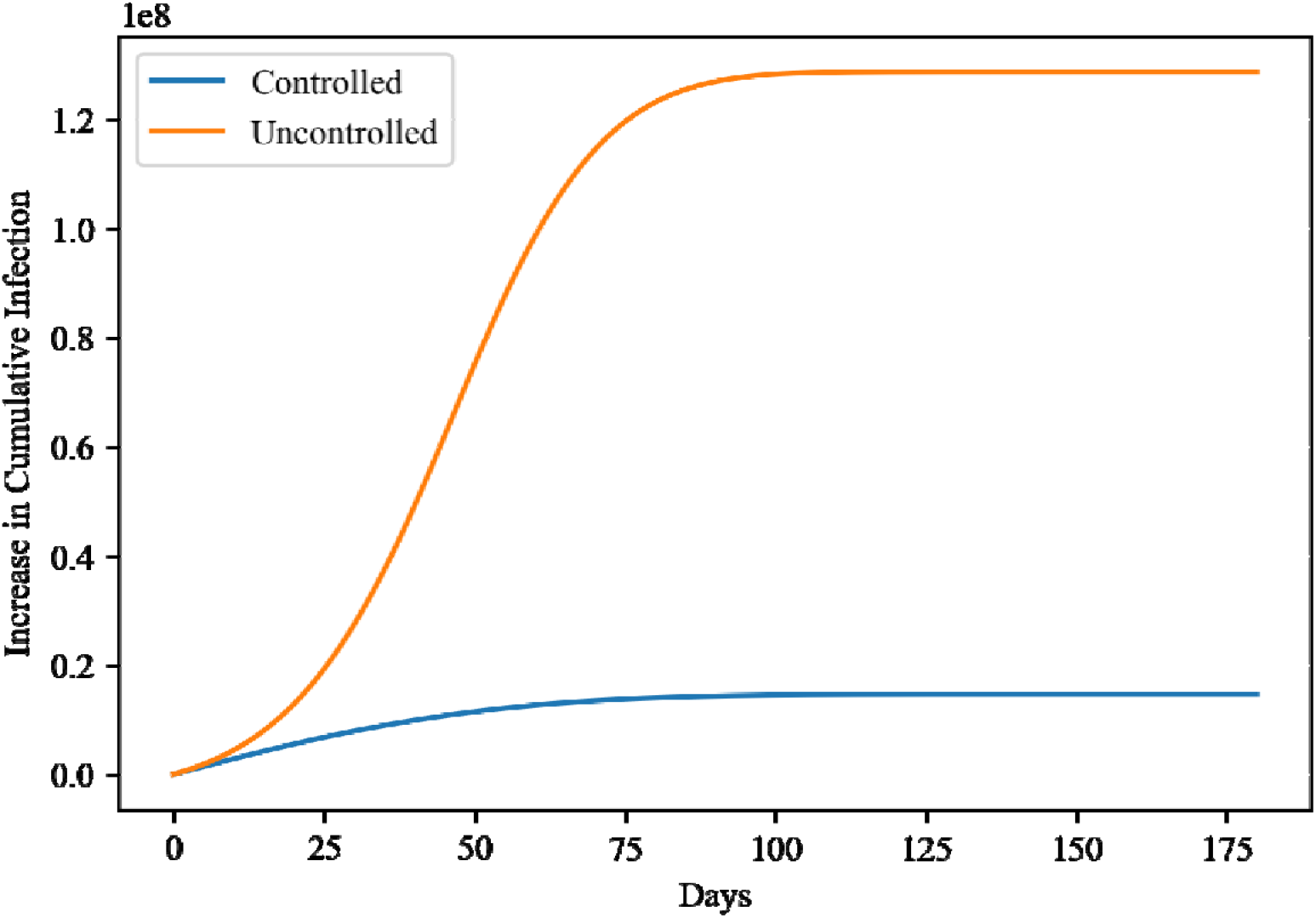
Increase in cumulative infection under Operation Warp Speed with vaccine compliance of 0.6, vaccine efficacy of 0.99, scenario 1 for non-pharmaceutical interventions (“controlled” case – blue) and scenario 6 consisting of no interventions (“uncontrolled” case – orange).

## Results

The carrying capacities and growth rates as functions of vaccine compliance and efficacies for each vaccination plan are provided as supplementary online material S1 – S4. In this paper, we focus on the adjusted growth rate in Figures 5-6 for the two federal plans, six scenarios (including 5 non-pharmaceutical interventions), and by varying vaccine efficacy as well as compliance. This allows to examine the synergistic effects of non-pharmaceutical interventions with vaccines while comprehensively accounting for key unknowns.

**Figure 6.**
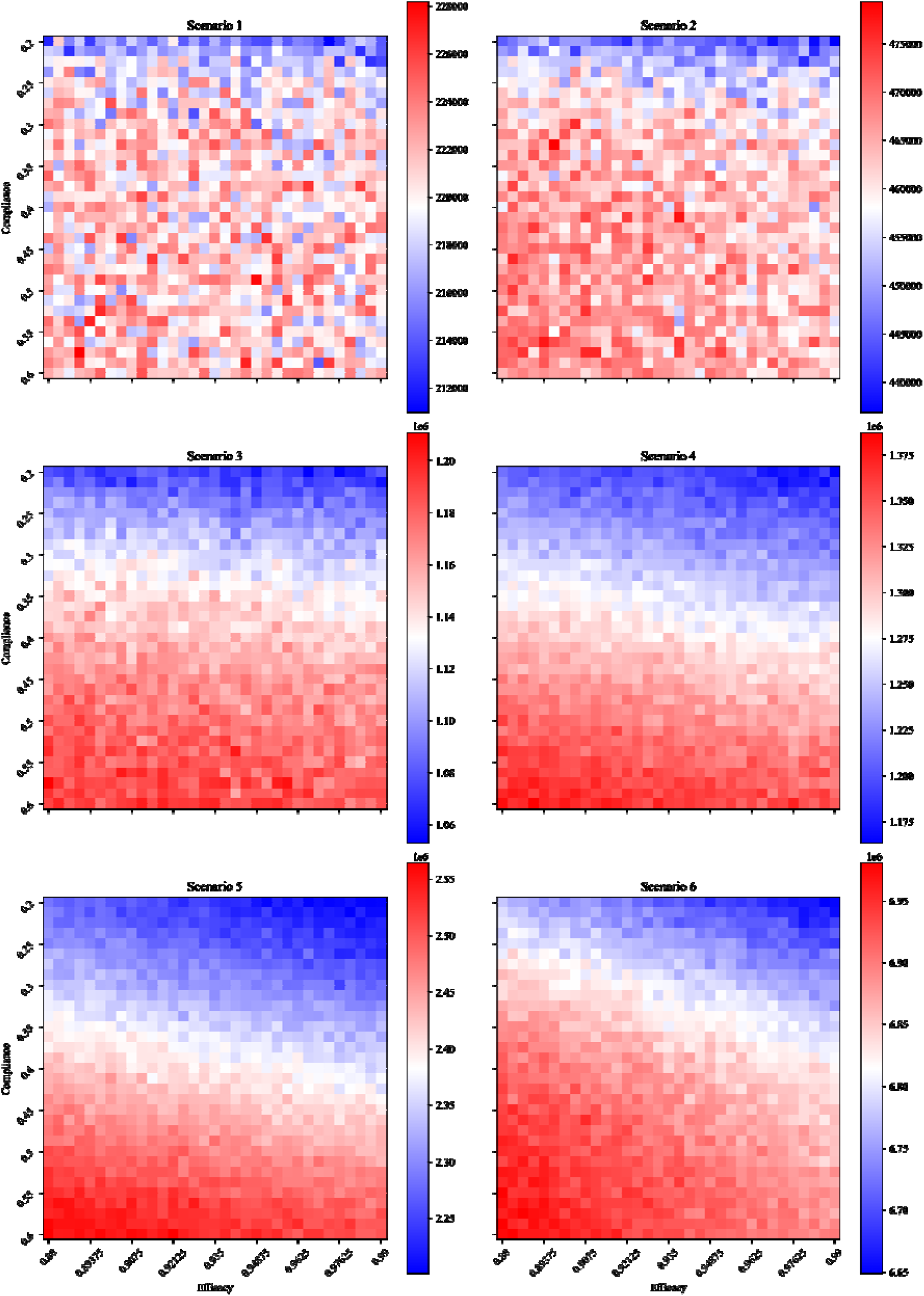
Adjusted growth rate (number of infected individuals on the worst day) as functions of vaccine compliance and efficacy under the Biden vaccination plan.

**Figure 7.**
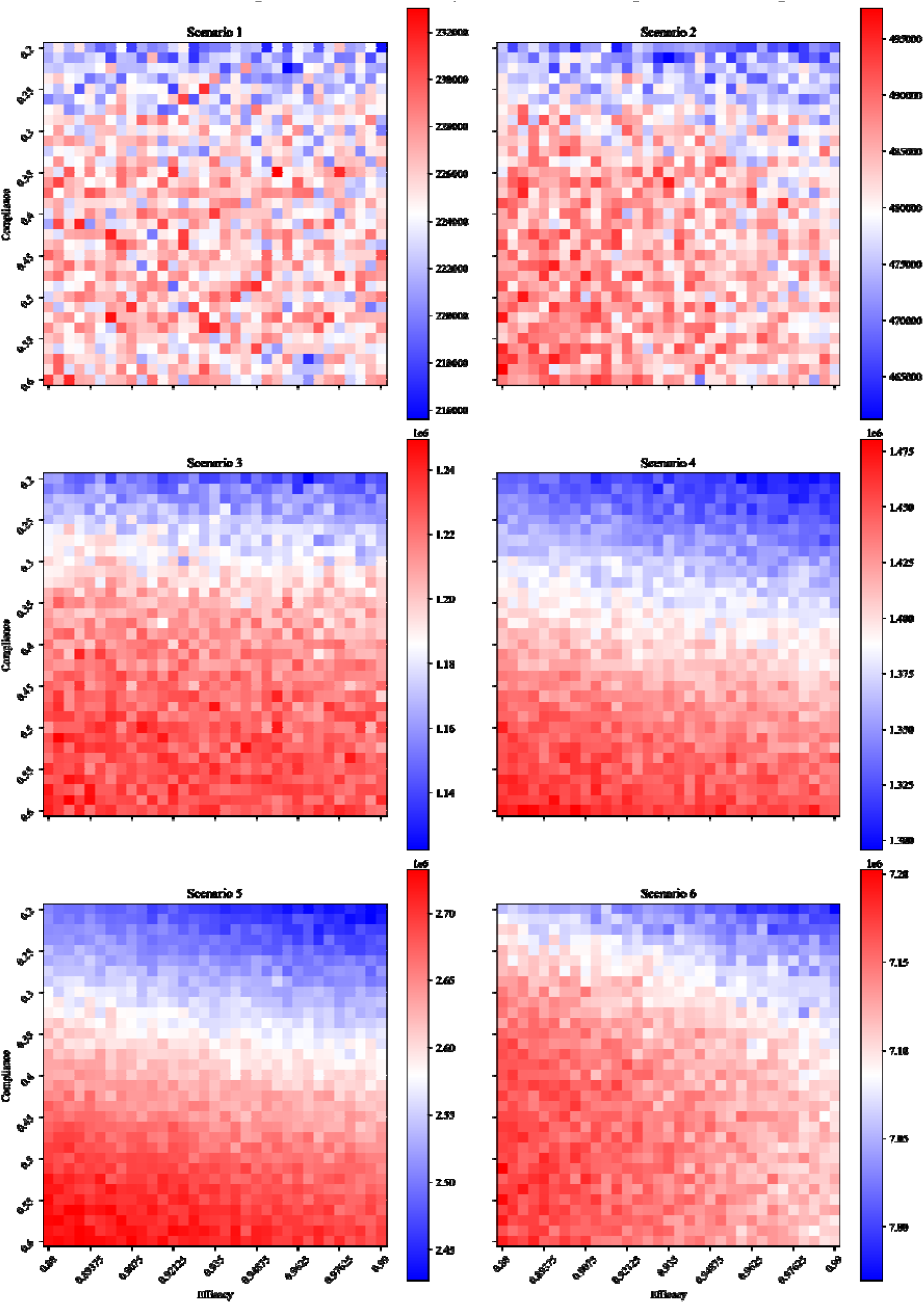
Adjusted growth rate (number of infected individuals on the worst day) as functions of vaccine compliance and efficacy under the Trump vaccination plan.

In comparing the two federal plans, the Biden plan showed more potency at controlling the infection across all intervention scenarios than the plan created under the previous administration. We note that even if a small fraction of the population seeks vaccines, and even if vaccines are less effective than announced, the vaccination campaign can reduce the total number of infections. Note that increasing the efficacies of vaccines results in lower infections for all scenarios and vaccine plans. This agrees with expectation since in our simulations, agents are not re-vaccinated upon having no immune response. Therefore, while holding all else equal, increasing the vaccine efficacy accelerates the growth of the immune population, thereby reaching herd immunity more quickly. In contrast, the dependence on compliance is much less intuitive and even leads to unintended consequences.

Typically, we assume that higher vaccine compliance will lead to lower overall infections, since the proportion of the immune population is upper bounded by the compliance. However, in both vaccination plans, only scenarios 1 and 2 yielded such results. For the rest of the scenarios (3 to 6), the dependence on vaccine compliance is apparently reversed, with some hinting towards a non-monotonic relationship (scenario 4 of the Biden plan and scenario 5 of the federal plan, for example). The reason behind this puzzling behavior is a combination of three factors: (i) vaccines are strictly administered in decreasing order of age, (ii) the elderly are going neither to work nor to school, hence they have fewer social ties than other age groups, which reduces their impact on preventing the spread of infections once immunized, and (iii) relative to the growth of infections in the scenarios in which the anomaly happen, the vaccine availabilities are too low.

As the vaccine compliance increases uniformly in the population, it means that a larger number of elderly in particular will seek vaccines. Given the vaccination strategy that focuses on older individuals, it will therefore take more time before anyone from the more connected/younger age groups can be vaccinated. During this time, the virus can continue to spread among the younger population, particularly because (i) the scenarios with counter-intuitive results (3 to 6) are among the least restrictive in terms of non-pharmaceutical interventions and (ii) elderly have a lower contribution to the spread of infections due to their more limited social ties. Therefore, while the elderly population will be better protected, the longer delay for the rest of the population means that by the times they are eligible for vaccinations, the infection has already spread, leading to overall higher infections.

This argument is most vividly illustrated by our animations in supplementary material S5 and S6, in which the distributions of the infected and immune population are plotted at each time step. These animations use showcase the no-intervention scenario (scenario 6) and Operation Warp Speed for the monthly vaccination capacity. Apart from the compliance, every other parameter including the random seed is fixed to be the same. Particular attention should be paid to the spread of infection among the elderly agents (i.e., over 65), as it most directly corroborates the reasoning above.

## Discussion

### Principal Results

The incoming CDC director predicted half a million death by mid-February [57], thus stressing the urgency of vaccination. However, vaccination is an unprecedented and complex endeavor whose success depends on many other variables such as vaccine compliance, vaccine efficacy, and the ongoing presence of non-pharmaceutical interventions. In line with expectations, our large-scale agent-based simulations show that vaccination can reduce the total number of infections across all possible scenarios. The capacity pledged under the new Biden plan (one million doses a day) would have a greater impact than the plan of the previous administration (“Operation Warp Speed”) when accounting for its initial delays.

Two key findings of our study are as follows. First, we demonstrate the necessity to maintain non-pharmaceutical interventions over the next six months. As interventions are relaxed (from scenario 1 offering the most control to scenario 6 offering no control), there is an increase in case count such that a return to normalcy is not achieved through vaccination but rather through a very high number of infected individuals. Second, there is an unexpected interplay between vaccination strategies, non-pharmaceutical interventions, and vaccination availabilities. As non-pharmaceutical interventions lose momentum (scenarios 3 and above), an increase in vaccine compliance leads to an unexpected increase in infections due in part on the low availability of vaccines and the priority on vaccinating elderly. More so than the observation that tighter non-pharmaceutical interventions result in the slower spread of infections, this result further delineates the necessity of preparing the population to continuing non-pharmaceutical interventions even as the vaccination progresses.

### Limitations

There are two main limitations to our current understanding of the COVID-19 pandemic and the vaccination campaign, which affect how our simulations account for (i) the number of vaccines that *can* be administered each month, and (ii) biological aspects.

First, an unprecedented vaccine campaign comes with logistical challenges and uncertainty given the complex array of factors involved. As a result, fewer than the expected number of doses may be administered: federal officials aimed at giving the first dose to 20 million people during December 2020, but various delays resulted in fewer than 3 million people receiving a first dose [58]. It was recently reported that “federal officials say they do not fully understand the cause of the delays” [58] and that the administration “pledged to immediately distribute millions of COVID-19 vaccine doses from a stockpile that the U.S. health secretary has since acknowledged does not exist” [59]. This situation has resulted in views that “much of the narrative earlier this year regarding Warp Speed’s preparation appears to be a sham” [60], reinforced by reports that the Biden administration found no vaccine distribution plan upon taking over from their predecessors [61]. Some of the factors causing a delay are known: there can be shipping delays or delays in administering doses due to a lack of hospital staff members as they are already caring for individuals infected with COVID-19. Other factors may be more surprising, such as the intentional destruction of vaccine doses by hospital staff [62]. As any simulation model is necessarily a simplification, we did not include factors whose value would be entirely unknown (e.g., what will be the shipment delay?) or whose existence is anecdotal given the total number of doses (e.g., intentional destruction or storage errors). We were limited in our ability to use real-world numbers on how many individuals received the vaccine as this data is captured at the state level and several states’ reporting systems have experienced errors [63]. Although there are efforts at centralizing data (e.g., national news outlets aggregate data across states [64]), the level and nature of errors differ across states, which is a challenge to estimate overall model uncertainty.

We have thus followed the federal plan for the number of individuals who can get vaccinated each month. Out of all the doses that are *planned*, fewer may be *distributed* and an even lower number may ultimately be *administered*. Our simulations are thus likely representing an upper bound on the number of vaccines administered, leading to *more optimistic results than in reality*. The gap is particularly pronounced in December and may remain significant in January, but early logistical issues and delays should be gradually addressed, such that the gap between federal expectations and actual implementation narrows over time.

Second, all biological aspects of the virus are based on the strains that dominated throughout 2020. Epidemiological studies from these strains have informed parameters such as transmissibility, incubation period, the proportion of asymptomatic carriers, the severity of symptoms and hence the course of the disease, as well as the efficacy of treatments or vaccines. The existence of different strains is well-established, as phylogenies have shown seven distinct lineages [refs 65, 66], but here has not yet been a documented need to ascribe different parameter values (i.e., different viral ‘behaviors’) to each strain. There are two possible reasons. First, there are relatively few mutations and thus a limited ‘chance’ of a drastically different outcome naturally occurring: the virus is “considered a slowly-evolving virus as it possesses an inherent proofreading mechanism to repair the mismatches during its replication” [ref 66]. Second, there has been little selective pressure on the virus, as it was spreading through a population that had never been exposed to an antigen (i.e., immunologically naïve). Both arguments are now changing.

A new strain from the lineage B.1.1.7, named Variant of Concern 202012/01 (denoted VOC-202012/01), emerged with an unusually large number of 23 changes in its genomes (including mutations and deletions) [ref 67]. Some of the biological changes make it easier for the virus to attach to its targets and enter cells, which is captured through epidemiological indicators as increased transmissibility [68 – 69]. This is relevant for our study, as this more contagious COVID-19 strain has been spreading in the USA and may dominate by March [70]. To date, there is no peer-reviewed evidence of an impact on disease severity or vaccine efficacy over a large population sample, but the function for some of the mutated parts remains unknown (hence the possibility of an impact on severity) and early studies over 20 volunteers suggests that antibodies from vaccines are only one-third as effective on some variants [71]. In addition, vaccination means that the virus is no longer spreading through an immunologically naïve population, thus creating selective pressure for functional mutations which can help the virus adapt. Our *simulation results are thus optimistic* as they use a lower transmissibility than provided by the new strain and as we did not worsen any of the other parameters to account for possible selective pressure.

Finally, we note that our model is *built very specifically for the USA*. It would not be accurate when transposed to another country with minimal changes (e.g., only reducing the population size). For example, stark differences in vaccine rollout strategies exist between the UK and the USA, which would affect our simulations. In the USA, two doses of the same vaccine are normally administered, as the CDC stated that “mRNA COVID-19 vaccines are **not** interchangeable with each other or with other COVID-19 vaccine products” [72]. However, new guidance from the UK allows a mix-and-match vaccine regimen in which the second dose may be from a *different* vaccine in exceptional circumstances (e.g., if the vaccine from the first dose is not available upon the patient’s return), even though clinical trials for mixed regimens are only due to be conducted at a later, unspecified time [73]. Another difference is that the UK frontloads the vaccine by delivering *as many first doses* as possible, which thus (*i*) no longer guarantees that a patient can receive the corresponding second dose upon return (hence raising the need for a mix-and-match) and (ii) potentially delays the delay before a second dose up to 12 weeks [73]. In contrast, the USA is against delaying the second dose [74], thus our model operates on the assumption that a patient can complete treatment on time.

## Supporting information

Supplementary Animation 1

Supplementary Animation 2

## Data Availability

This study uses secondary data and models which have been publicly released.

## Acknowledgements

The authors thank the Microsoft AI for Health program for supporting this research effort through a philanthropic grant. The sponsor did not influence the design, methods, or analyses in this study.

## Abbreviations

ABM: An Agent-Based Model is a computational model that simulates how individuals (i.e., ‘agents’) change through interactions with others and their environment.
CDC: The Centers for Disease Control and Prevention is a national public health institute in the United States.
COVID-19: Disease caused in humans by a new strain of the severe acute respiratory syndrome coronavirus 2, SARS-CoV-2 (initially named 2019-nCoV).
VOC-202012/01: Variant of Concern 2020/12/01 is a new strain, first identified in the UK and currently spreading in the USA, whose unusually large number of changes in genomes result in higher transmissibility.

