## Supplementary figures and images for "Returning to a normal life via COVID-19 vaccines in the USA: a large-scale agent-based simulation study"

### Supplementary Figure 1

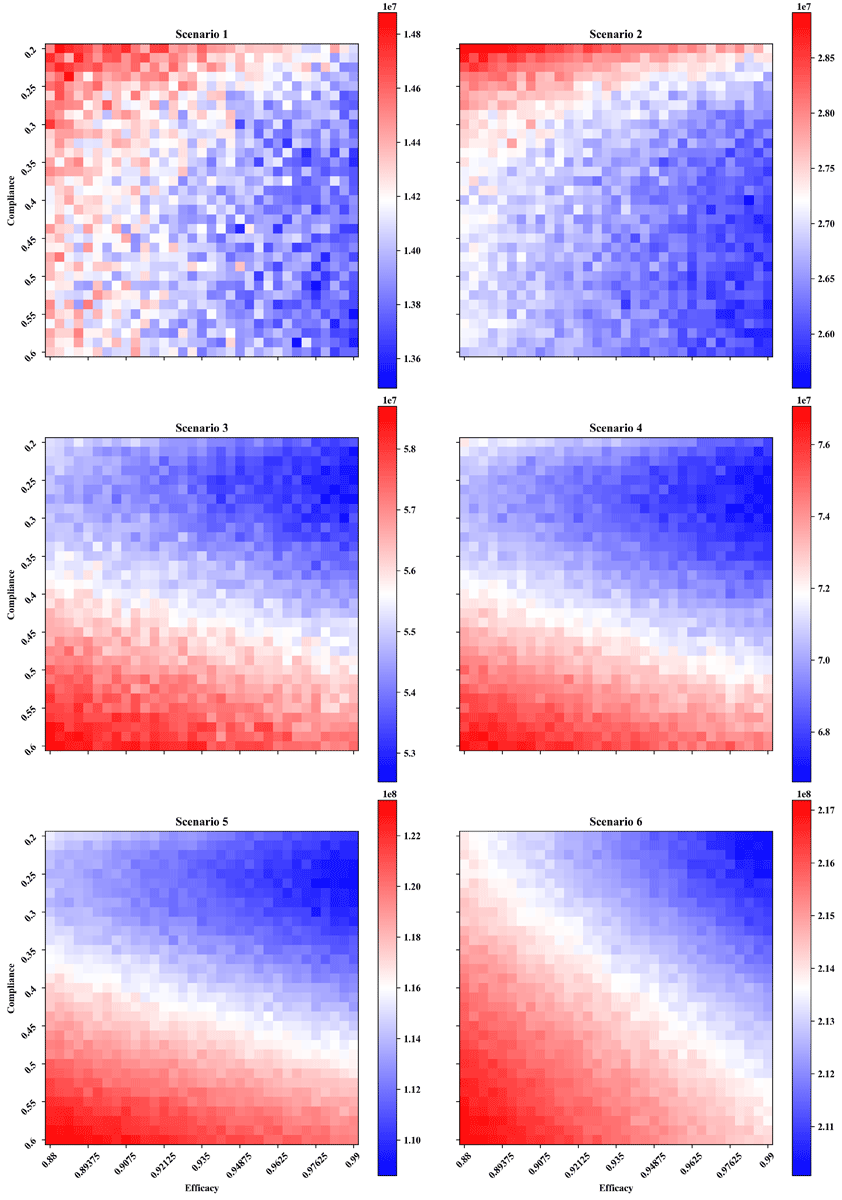

### Supplementary Figure 2

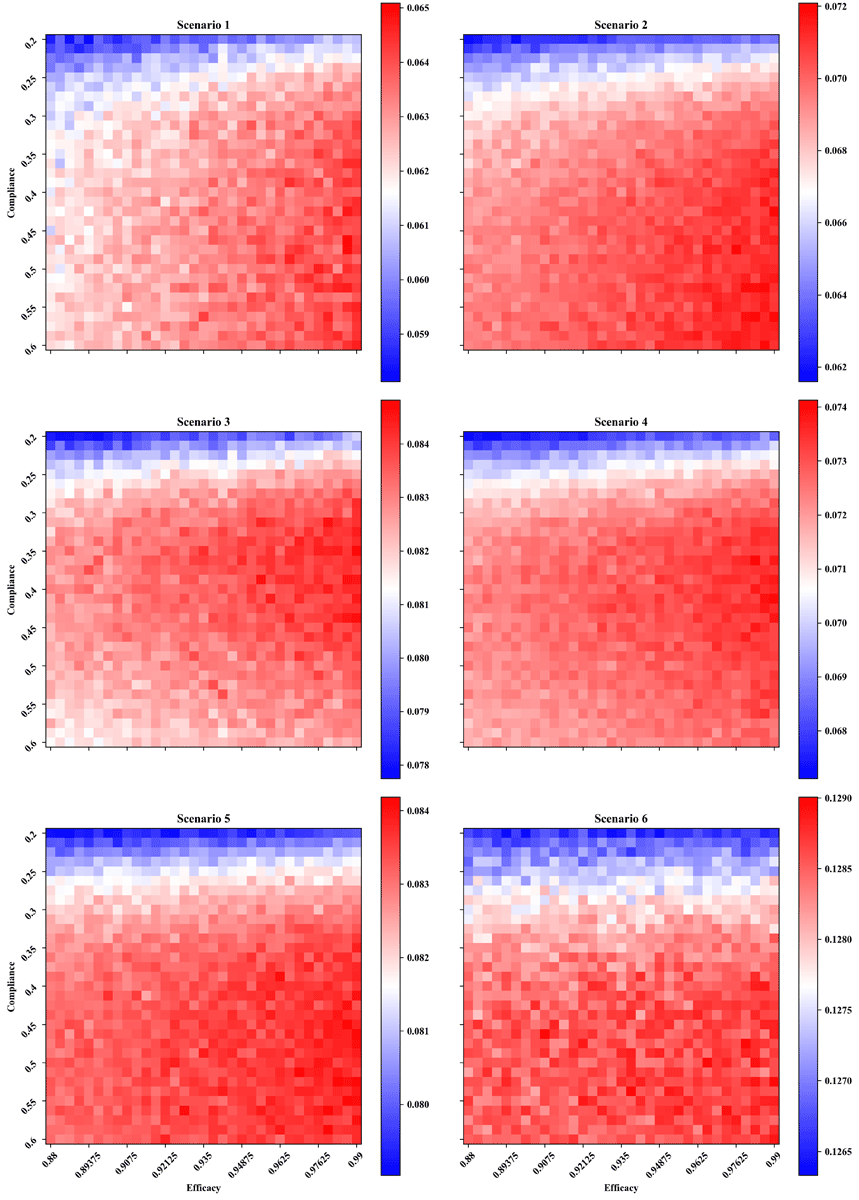

### Supplementary Figure 3

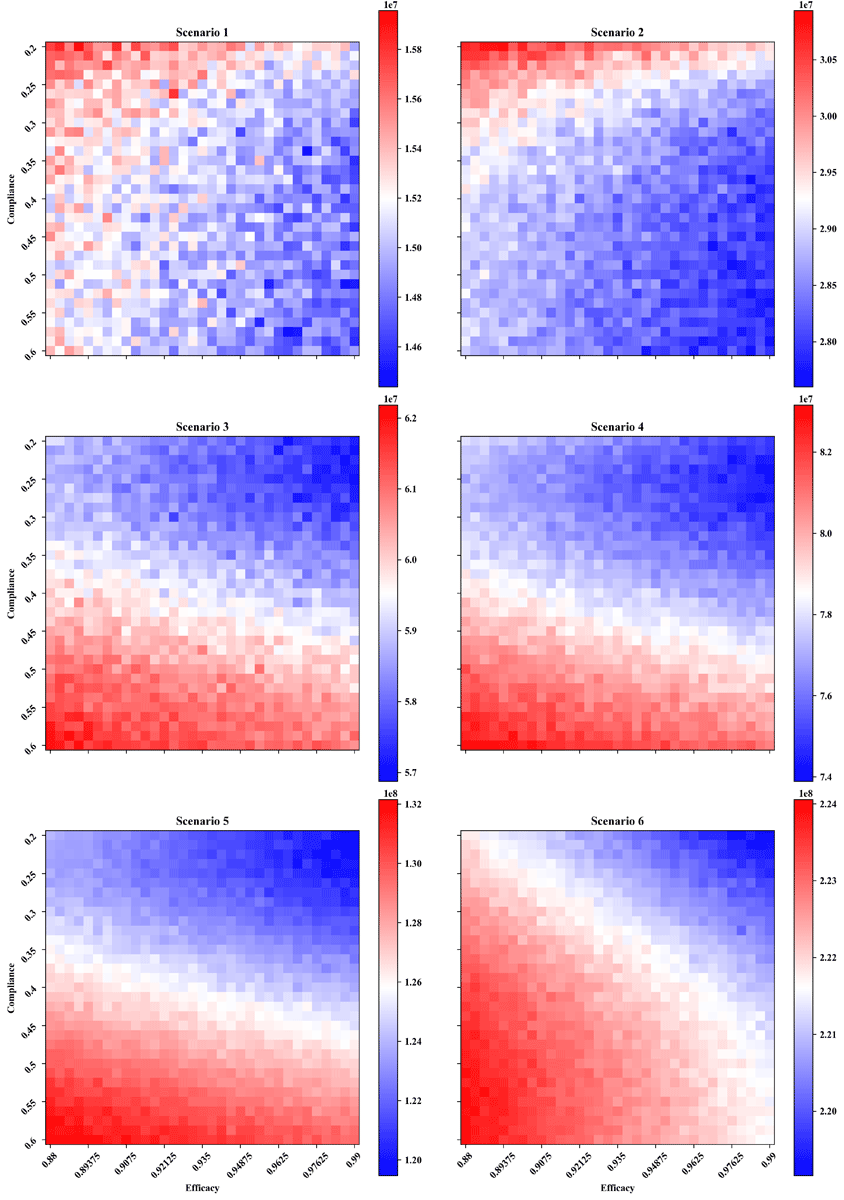

### Supplementary Figure 4

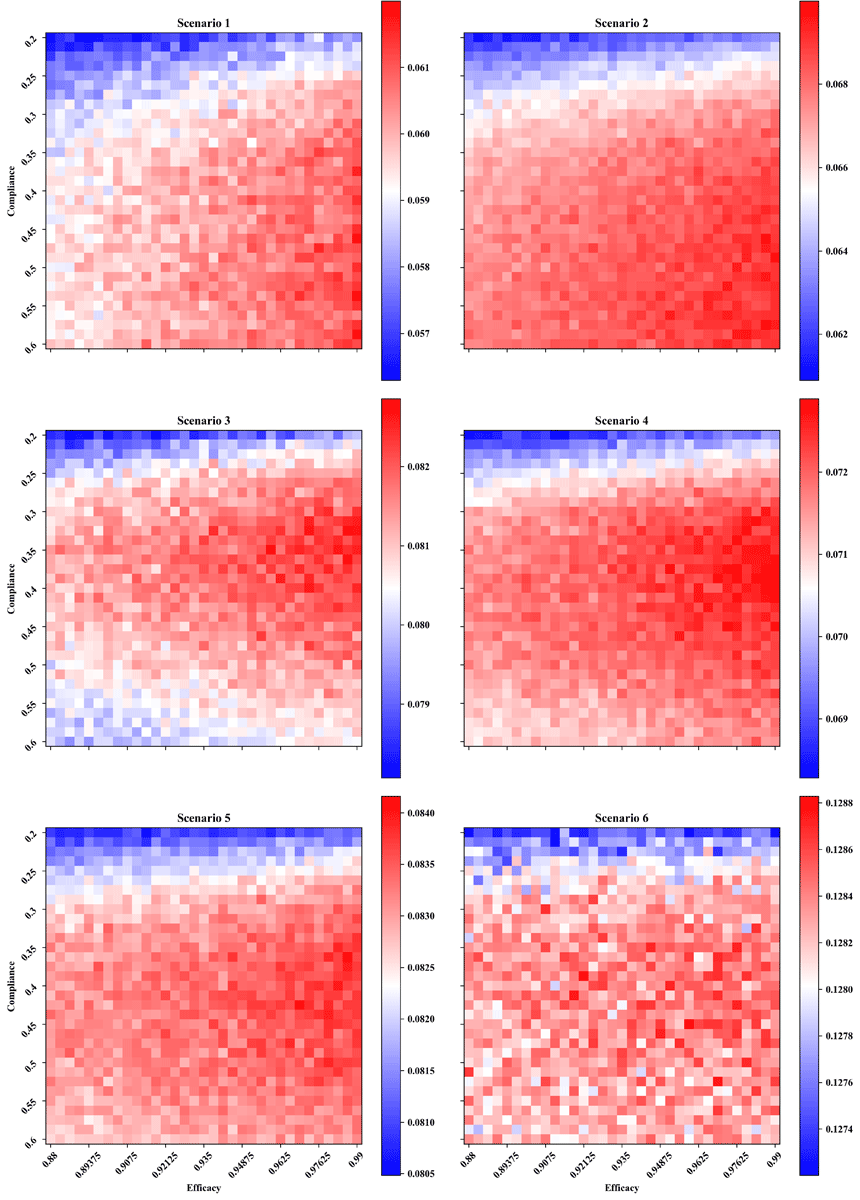
